# Rare Variants in Inborn Errors of Immunity Genes Associated with Covid-19 Severity

**DOI:** 10.1101/2022.03.09.22270766

**Authors:** Panhong Liu, Mingyan Fang, Yuxue Luo, Fang Zheng, Yan Jin, Fanjun Cheng, Huanhuan Zhu, Xin Jin

## Abstract

Covid-19 is a contagious disease caused by SARS-CoV-2, a novel severe acute respiratory syndrome coronavirus. Common variants and networks underlying host genetic mechanisms have been extensively studied to identify disease-associated genetic factors. However, there are few studies about the rare variants, typically inborn errors of immunity, in understanding the host genetics behind Covid-19 infection, especially in the Chinese population. To fill this gap, we investigate likely-deleterious missense and high-confidence predicted loss-of-function variants by (a) performing gene- and pathway-level association analyses, (b) examining known genes involved in type I interferon signaling and others previously reported in Covid-19 disease, and (c) identifying candidate genes with accumulating mutations and their potential protein-protein interactions with known genes. Based on our analyses, several putative genes and pathways are uncovered and worth further investigation, for example, genes *IL12RB1, TBK1*, and *TLR3*, and pathways Tuberculosis (hsa:05152), Primary Immunodeficiency (hsa:05340), and Influenza A (hsa:05164). These regions generally play an essential role in regulating antiviral innate immunity responses to foreign pathogens and in responding to many inflammatory diseases. We believe that to some extent, as an acute inflammatory disease, Covid-19 is also affected by these inborn errors of immunity. We hope that the identification of these rare genetic factors will provide new insights into the genetic architecture of Covid-19.

## Introduction

Since the December of 2019, the coronavirus diseases 2019 (Covid-19) (Gorbalenya et al. 2020) caused by the SARS-CoV-2 virus (Severe Acute Respiratory Syndrome Coronavirus 2) (N. Zhu et al. 2020) has spread rapidly across the world. By January 2022, the ongoing SARS-CoV-2 pandemic has caused more than three hundred and sixty million confirmed cases and more than five million deaths. Host genetic factors have been shown to play critical roles in the disease susceptibility and severity. The Covid-19 Host Genetics Initiative (Covid-19 HGI, https://www.covid19hg.org/) is an international initiative to share the results of host genome-wide associations study (GWAS) meta-analysis of Covid-19 disease. The most recent Covid-19 HGI release 6 has reported 24 lead SNPs (P < 5e-8) mapped to nearly 136 genes, such as *LZTFL1, ABO, OAS1, DPP9, IFNAR2* (Initiative and Ganna 2021). The estimated heritability of Covid-19 symptoms explained by these common variants was 6.5% (Pairo-Castineira et al. 2021). A twin study with participants from the TwinsUK cohort reported that 31% of phenotypic variance of predicted Covid-19 is due to host genetic factors (Williams et al. 2020). This leads to a large proportion of unexplained heritability (nearly 25%), commonly referred to as “missing heritability”. There is increasing evidence that rare variants also make a major contribution to missing heritability of many complex diseases and traits (Zuk et al. 2014; Hunt et al. 2013; Misawa et al. 2020).

Recently, the rare variants attracted researchers’ attention in elucidating the missing heritability of Covid-19 susceptibility and severity. For example, Zhang et al. found that the rare predicted loss-of-function (pLoF) variants in the IRF7- and TLR3-dependent type I interferon (IFN) pathway were enriched in patients who developed risky Covid-19 (Q. Zhang et al. 2020). Smieszek et al. reported that pLoF variant in gene *IFNAR2* (c.966C>A/p.Y322X) might play a role not only in clinical manifestation of Covid-19 but also in the response to vaccination (Smieszek et al. 2021). In addition, Mantovani et al. found that pLoF variants in *TLR7* (c.3094G>A/p.A1032T, c.901T>C/p.S301P) occurred in severely affected male patients and downregulated the expression of TLR7 pathway (Mantovani et al. 2021). As previously reported, the rare variants were more likely to be functional and tended to have stronger effects on complex diseases (Gorlov et al. 2011). The study of genetic effects of rare variants is necessary to elucidate the severity of Covid-19.

To explore the genetic contributions of rare variants in Covid-19 patients with inborn errors of immunity, we recruited and investigated nearly 500 hospitalized patients from Union hospital of Tongji Medical College of Huazhong University of Science and Technology (*abbr*. Union hospital) (H. Zhu et al. 2021). Based on patients’ genomic data and clinical information, we carried out three major analyses to investigate the effects of host rare variants: (a) gene- and pathway-level tests of these rare variants between *severe* and *non-severe* patients; (b) examination of the significance of previously reported rare variants and genes in our dataset; and c) rare mutation accumulation analysis and protein-protein interaction (PPI) network analysis in only *severe* patients. From these analyses, we (a) identified candidate functional pathways that are responsible for innate immune disorders and respiratory diseases, such as Tuberculosis (hsa:05152), Primary Immunodeficiency (hsa:05340), and Influenza A (hsa:05164); (b) successfully replicated two Covid-19 associated SNPs (rs780744847 and rs541048548) mapped on genes *TLR3* and *ICAM3*, respectively; and (c) suggested several candidate genes, including *IL12RB1, TBK1*, and *TLR3*, which might be involved in SARS-CoV-2 cell entry, host immune responses, and Covid-19 disease severity.

Until now, literatures based on Chinese population have replicated and discovered some Covid-19-associated common variants (Wang et al. 2020; H. Zhu et al. 2021; P. Wu, Chen, et al. 2021; P. Wu, Ding, et al. 2021), but genetic background of rare variants is currently insufficiently understood in Chinese population. Our work is an effort to fill this gap. We hope that it will serve as useful scientific reference to assess the genetic mechanism of rare variants in Covid-19 and advance our understanding of disease etiology.

## Materials and Methods

### Patient Recruitment and Quality Control

All subjects in this study were collected from the Union hospital. We used PLINK 2.0 (Chang et al. 2015) to infer sex of individuals from SNP genotypes and VerifyBamID (F. Zhang et al. 2020) to assess the level of DNA contamination. Individuals were excluded if their PLINK inferred sex was inconsistent with that of clinical recorded. We also removed individuals with estimated contamination rates greater than 0.05. After sample quality control, there were 451 unrelated individuals with 159 mild and moderate patients, and 292 severe and critical patients. The severity classification criteria were made by the National Health Commission of P.R. China (Z. Wu and McGoogan 2020). We reclassified the mild/moderate patients as *non-severe* patients and the severe/critical patients as *severe* patients.

### Genotype Calling

The blood samples of some patients were collected at different time points during hospitalization. To increase the average depth of study, sequence fastq files of each patient were merged together to generate one GVCF file by BWA and Sentieon Genomics software (Freed et al. 2017). Joint variant calling was then performed on GVCF files of all participants using the Sentieon GVCFtyper algorithm. The resulting VCF file was used for subsequent genomic analyses. After the application of excessHet (<54.69) filter, Variant Quality Score Recalibration (VQSR) was completed by using the Genome Analysis Toolkit (GATK version 4.1.2) (DePristo et al. 2011). To improve the genotyping accuracy, we used the Beagle 4.0 software (Browning and Browning 2016) to perform LD-based genotype refinement by taking genotype likelihoods as inputs. Low-quality variants with dosage imputation score R2 < 0.3 by Beagle4.0 were filtered out.

### Principal Component Analysis

Principal component analysis (PCA) was performed using a subset of autosomal bi-allelic SNPs by applying PLINK 2.0 (Chang et al. 2015). Several restrictions were applied to select SNPs for PCA analysis, including keeping SNPs with minor allele frequency (MAF) ≥ 5%, Hardy–Weinberg Equilibrium P ≥ 1e-6, and removing one of a pair of SNPs if the LD was greater than 0.5 (in a window of fifty SNPs with a stop of five SNPs).

### Functional Annotation

We annotated rare variants (MAF < 0.5%) in our final call set by using the Ensembl Variant Effect Predictor (VEP, build 103, GRCh38) (McLaren et al. 2016) with default parameters. The databases for annotation included dbSNP (Sherry et al. 2001), gnomAD (Karczewski et al. 2020), and 1000 Genomes Project (Clarke et al. 2012). In addition, we used Combined Annotation Dependent Depletion (CADD) score to predict missense variants that had potential effects on protein function. The CADD score was annotated by CADD plug-in (Kircher et al. 2014). Missense variants with CADD score > MSC (Mutation Significance Cut-off) score (95% confidence interval) (Itan et al. 2016) were predicted as likely-deleterious missense variants. We also used LOFTEE (Karczewski et al. 2020) plugin to identify high-confidence pLoF (HC-pLoF) for stop-gained, frameshift, and splice site disrupting variants. Finally, we focused on the likely-deleterious missense and HC-pLoF variants in the subsequent analyses.

### Rare Variants Analyses

To investigate the cumulative effects of multiple rare variants, we performed gene-based association analysis using KGGSeq 1.0 (Li et al. 2017) with the sequence kernel association test (SKAT) (M. C. Wu et al. 2010), the Optimized SKAT (SKAT-O) (Lee, Wu, and Lin 2012), and Burden test. We further carried out pathway-based analysis by testing the Kyoto Encyclopedia of Genes and Genomes (KEGG) gene sets (Kanehisa and Goto 2000). The adjusted covariates included age, sex, and the top six principal components. The gene-based and pathway-based p-values were then multiple testing corrected by Benjamini-Hochberg method. We defined the suggestive significance threshold for gene-based association test as 1e–6 and for pathway-based association test as 0.05.

We also focused on 13 type I IFN genes (denoted as IFN-genes) that were found an enrichment in life-threatening Covid-19 study (Q. Zhang et al. 2020) and 136 genes located in 50kb of risk lead SNPs reported by the Covid-19 Host Genetics Initiative (release 6, denoted as HGI-genes) (Initiative and Ganna 2021). The mutation accuracy of variants in these 148 candidate genes (one overlap between 13 IFN-genes and 136 HGI-genes) was manually checked by using Samtools 1.10 (Danecek et al. 2021).

Finally, we performed an analysis of rare variant accumulation in genes identified by two approaches. The first approach detected genes if there was one variant met the following two conditions: (a) the mutations occurred in only *severe* patients, and (b) the variant harbored no less than three effect allele counts. We denoted these genes as “individual variant-driven genes”. The second approach determined genes if (a) all mutations in the gene occurred in only *severe* patients, and (b) the total number of mutations in the gene is at least three. We denoted these genes as “all variant-driven genes”. We note that, genes identified by the two methods may have some overlaps. Each of the two gene sets was then used for network analysis of protein-protein interactions (PPI) with the above 148 known candidate genes. We used the STRING version 10.5 (Search Tool for the Retrieval of Interacting Genes/Proteins) (Szklarczyk et al. 2019) to build the PPI network. The minimum required interaction score to highest confidence was set to 0.900.

## Result

### Participant Characteristics

In this study, participants included 451 Covid-19 patients aged from 23 to 97 years old and all declared Han Chinese population. In Table 1, we provided participant characteristics for *non-severe* and *severe* patients, respectively. A total of 159 (35.25%), 292 (64.75%) patients were grouped as *non-severe* and *severe*, respectively. The same as previously reported (Cummings et al. 2020; Yang et al. 2020), older age (*severe*: an average of 64 y/o vs. *non-severe*: an average of 58 y/o, t-test *p* = 4.6e-05) and male sex (*severe* 52.74% vs. *non-severe* 42.14%, Fisher’s exact test *p* = 0.04) were at a higher risk of developing severe symptoms.

**Table 1.** Participant Characteristics.

|  | Number, n (%) | Male gender,<br>n (%) | Age, average<br>(sd) | Depth,<br>average (sd) |
| --- | --- | --- | --- | --- |
| All patients | 451 |  |  | 21.67 |
| Non-Severe | 159 (35.25%) | 67 (42.14%) | 58.33 (14.62) | 19.04 (8.94) |
| Severe | 292 (64.75%) | 154 (52.74%) | 64.11 (13.31) | 23.1 (10.43) |

### Data Quality

After quality control, the dataset consisted of 22,107,585 and 680,522 variants from autosomes and X chromosome, respectively (Figure 1). Then we compared chip array sequencing results with genotype after LD-based refinement by Beagle 4.0 on 218 individuals. The heterozygote concordance rate increased from an average of 94.4% to 97.4%, and the improvement is more dramatic for samples with lower sequencing depth (Figure 2A). After filtering in variants by imputation score DR2 > 0.3, the final dataset for further analyses had a total of 22,532,360 variants, and the PCA on 575,888 autosome SNPs detected no outlier samples (Figure 2B).

**Figure 1.**
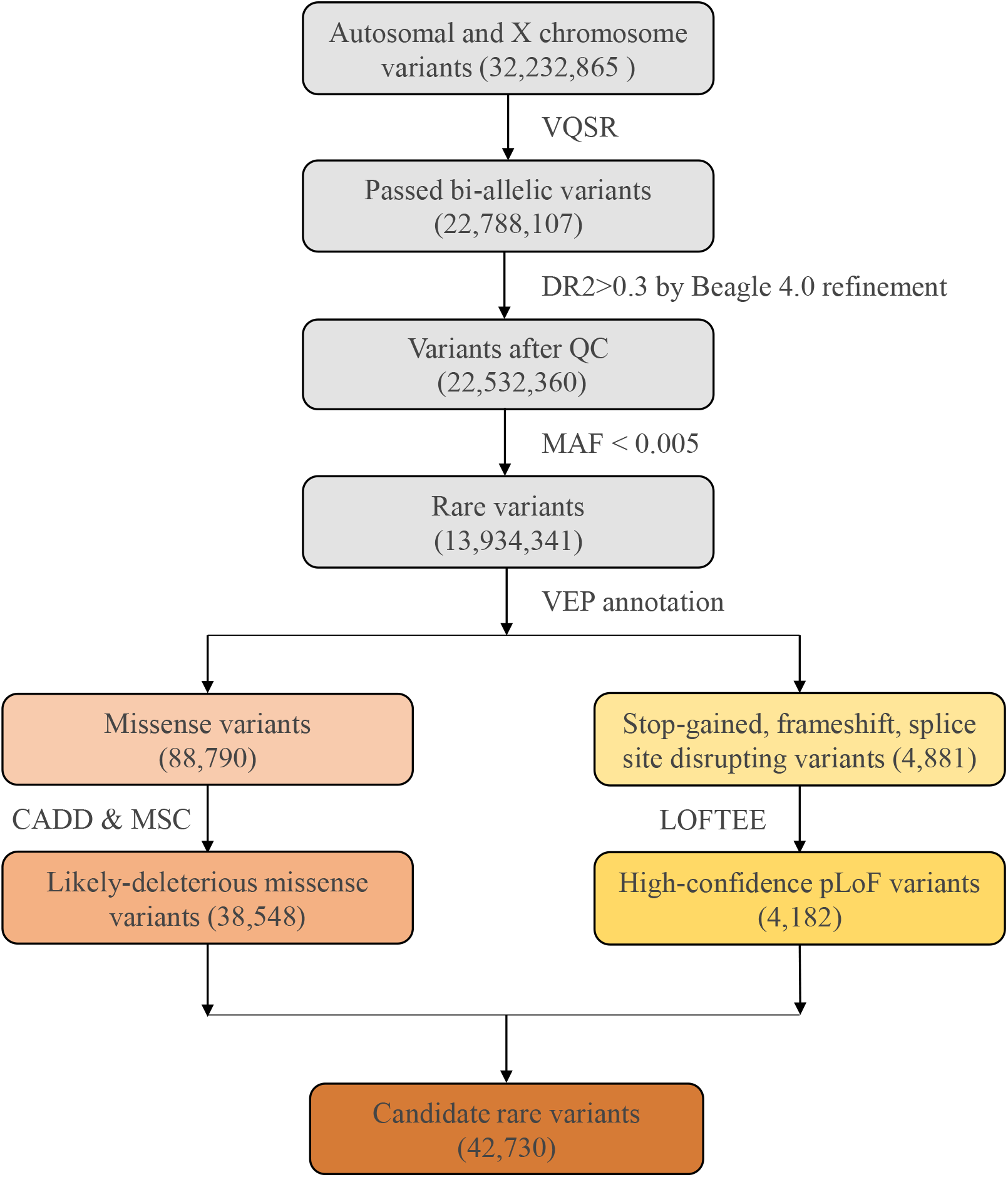
The flow diagram of rare variants analysis. A total of 32,232,865 variants were identified from the 451 Covid-19 patients with whole genome sequencing. After filtering by VQSR and MAF, 13,934,341 rare variants were annotated by VEP, and 42,730 candidate variants were included.

**Figure 2.**
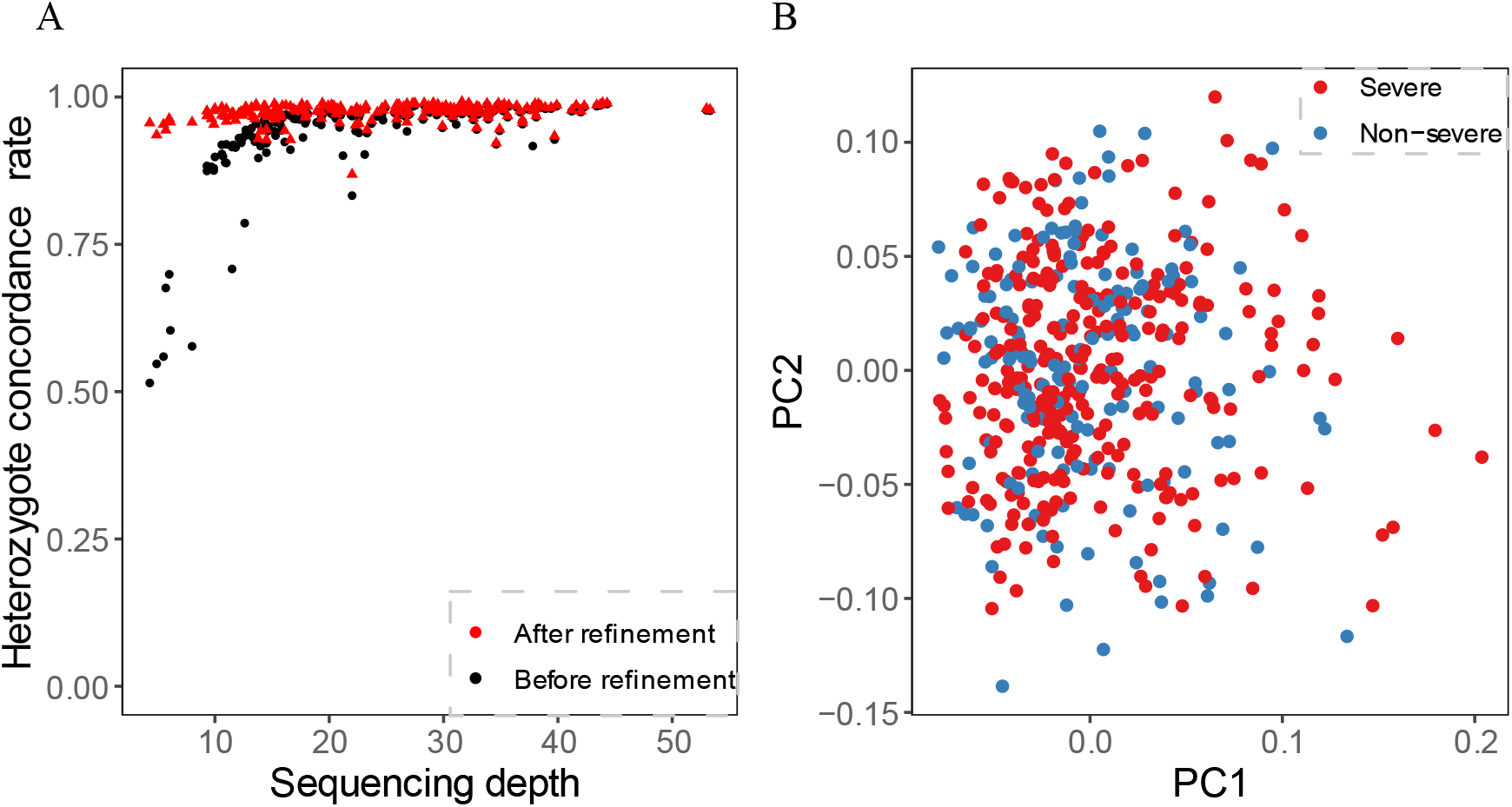
Quality estimate of the cohort. (A). Heterozygote concordance rate versus sequencing depth for 218 array-genotyped individuals. (B). PCA of 159 non-severe and 292 severe patients.

### Rare Variants Statistics

After filtering by MAF, we obtained a total of 13,934,341 rare variants for VEP annotation. Among the resulting annotations, there were 88,790 missense variants and 4,881 pLoF variation (including stop-gained, frameshift, and splice site disrupting variants). Damaging effects of these missense and pLoF variants were then predicted by CADD and LOFTEE plug-in, respectively. About 43.41% missense variants were predicted as likely-deleterious missense variants (38,548) and 85.68% pLoF variants were predicted as HC-pLoF variants (4,182). Thus, in total, 42,730 predicted likely damaging variants were applied for further analysis. For both likely-deleterious missense variants and HC-pLoF variants, we first tested the difference of their numbers between *non-severe* and *severe* patients and found no significant difference (Supplementary Table S1).

### Gene-and Pathway-level Analysis of Rare Variants

The gene-level analysis of rare variants was performed between *severe* and *non-severe* patients via KGGSeq. We performed the gene-based tests for genes mapped by all rare variants, likely-deleterious missense variants, and HC-pLoF variants, respectively. The gene-based analyses did not identify putative genes that passed the significance threshold of 1e-6 (Supplementary Figure S1A-C).

Furthermore, we leveraged the biological knowledge that sets of genes acting together in pathways. In total, we tested 307 KEGG pathways and detected Tuberculosis (hsa:05152, p.adjust = 0.036) between *severe* and *non-severe* patients on likely-deleterious missense and HC-pLoF variants (Supplementary Table S2). Tuberculosis (TB) is an airborne infectious disease caused by Mycobacterium tuberculosis (Mtb). It first attacks the lungs, then other parts of the body through circulatory system. This transmission characteristics is very similar to that of Covid-19. As previously reported, Tuberculosis pathway was significant with acute respiratory distress syndrome and lung injury in mice and human (Sweeney et al. 2017). The TB/Covid-19 Global Study Group observed a phenomenon that TB and SARS-CoV-2 might be co-infected, i.e., TB was often diagnosed concurrently or after Covid-19 infection and the co-infection might account for increased case fatality rate (Group 2021). Our finding brought up a possible explanation that patients with rare mutations enriched in Tuberculosis pathway were more likely to develop severe Covid-19 symptoms. When focused on only pLOF variants enriched on KEGG, two significant pathways highlighted: Primary immunodeficiency (hsa:05340, p.adjust = 0.014) and Influenza A (hsa:05164, p.adjust = 0.021) (Supplementary Table S2). Primary immunodeficiencies (PID) are a group of potentially serious disorders that can cause increased susceptibility to severe infections, autoimmune diseases, and malignancy. Several studies revealed that patients with PID displayed higher morbidity and mortality from Covid-19 (Shields et al. 2021; Ho et al. 2021; Babaha and Rezaei 2020). Influenza is an infectious respiratory disease caused by influenza virus. Bibert et al. observed that gene pathways involved in the detection of Influenza A overlapped with those involved in the detection of SARS-CoV-2 virus (Bibert et al. 2021). In these two biological pathways, three functional genes, *IKBKG, IRF7*, and *IFNAR1*, were previously identified to have an effect on Covid-19 severity (Q. Zhang et al. 2020).

### Tested on 148 Candidate Genes

In addition to uncovering unknown possibly associated genes or pathways, we also tested 148 previously reported candidate genes, with 13 in the type I IFN pathway (Q. Zhang et al. 2020) and 136 located within 50kb of significant common variants in the Covid-19 HGI (Initiative and Ganna 2021). Specifically, we focused on likely-deleterious missense and HC-pLOF variants to aggregate potential effects of rare variants. For missense variants in both IFN-and HGI-genes, we did not detect significant difference between *severe* and *non-severe* patients. In the 13 IFN-genes, we found one HC-pLoF variant rs780744847 (c.1180C>T/p.R394*) on *TLR3* mutated in only *severe* patients but no mutations in *non-severe* patients. It was reported that the *TLR3* deficiency may lead to increased incidences of viral infections and impair the production of type I IFN throughout SARS-CoV-2 infection (Q. Zhang et al. 2020). Moreover, mutations of inborn errors of TLR3-dependent type I IFN immunity more often occurred in highly critical patients than in mild patients and healthy controls. For the 136 HGI-genes, we found that the number of HC-pLoF mutations occurred in *severe* group was more than that of *non-severe* groups (16 in *severe* and 2 in *non-severe* patients, Fisher’s exact test p = 0.043) (Table 2). We also detected a HC-pLoF variant rs541048548 (c.1053del/p.A352fs) on *ICAM3* only mutated in *severe* patients. The gene *ICAM3* played an important role in the immunopathogenesis of SARS virus (Chan et al. 2007) and had been reported that its expression was downregulated in asymptomatic Covid-19 cases compared with symptomatic patients (Masood et al. 2021).

**Table 2.**
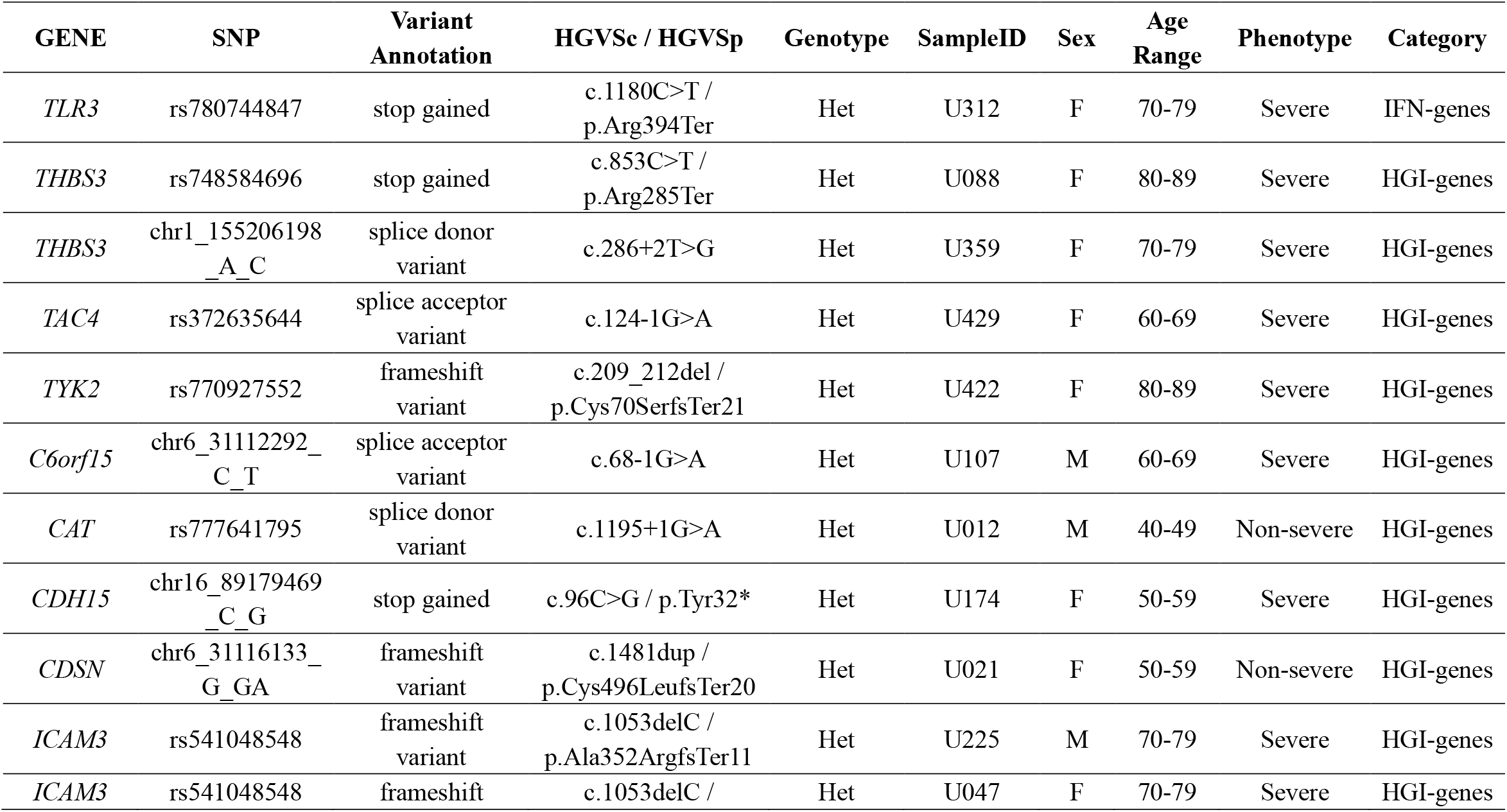
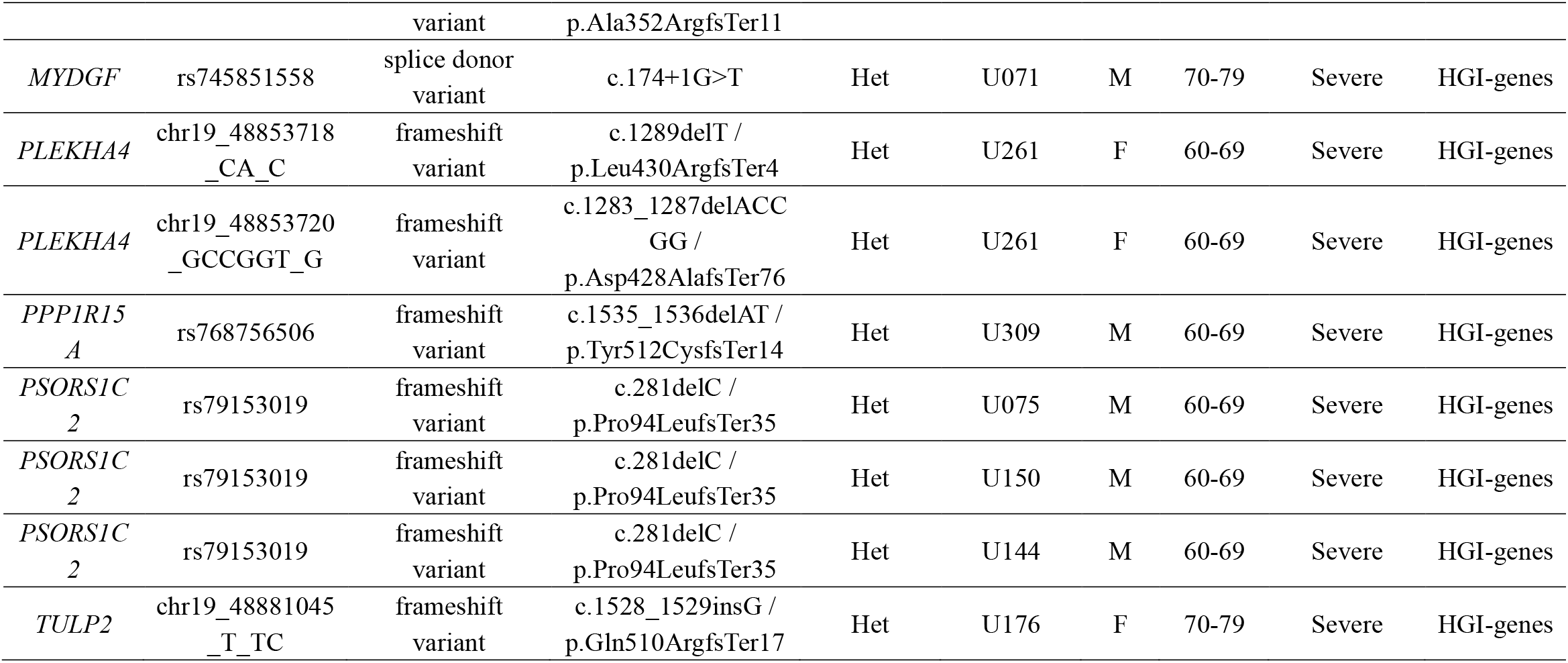
The pLoF variants identified in covid-19 patients in 148 candidate genes.

### Mutation Accumulation Analyss

In the mutation accumulation analysis, we first investigated whether there were potentially functional mutations unique to *severe* patients. We filtered in rare variants mutated in only *severe* patients and with minor allele count (MAC) greater than or equal to three. This resulted in 756 rare variants mapped to 700 genes. Among these variants, we observed a very rare mutation rs777044791 in gene *CCR3* at locus 3p21.31 (Table 3). The physical distance between rs777044791 and rs11385942 is 0.43MB (GRCh38), a distance typically flanked into the same genomic region (Casto and Feldman 2011). The variant rs11385942 is a common variant located at locus 3p21.31 and was first identified to be associated with respiratory failure due to Covid-19 from GWAS analysis in the Italian and Spanish population (Ellinghaus et al. 2020). This finding was repeated in other studies based on European populations (Ellinghaus et al. 2020; Pairo-Castineira et al. 2021; Shelton et al. 2021), verifying its effects on Covid-19 disease. In Chinese population, common variant studies at this locus did not replicate significance (P. Wu, Chen, et al. 2021; H. Zhu et al. 2021; Wang et al. 2020), and no rare variant studies had been conducted. Our work filled this gap and revealed 3p21.31 as the Covid-19 risk locus.

**Table 3.** The comparison of allele frequency for two loci.

|  | **rs777044791** | **rs11385942** |
| --- | --- | --- |
| CHROM | chr3 | chr3 |
| POS (hg38) | 46,266,186 | 45,834,967 |
| ALT | T | GA |
| REF | C | G |
| Variant Annotation | Missense Variant | Intron Variant |
| ***Allele frequency*** |  |  |
| Severe (N = 292) | 0.005 | 0 |
| Non-severe (N = 159) | 0 | 0 |
| ChinaMAP | 0.002 | 0.004 |
| 1000G_EAS | 0 | 0.005 |
| 1000G_EUR | 0 | 0.0805 |
| 1000G_SAS | 0 | 0.296 |
| 1000G_AFR | 0 | 0.053 |
| gnomAD_EAS | 0.0005 | 0.0006 |

Then, we performed PPI network analysis for the 700 “individual variant-driven” genes with the 148 known genes (Figure 3A). From the results, we found two candidate genes *IL12RB1* and *TRAF3IP3* that had extensive interactions with IFN-and HGI-genes. Gene *IL12RB1* (Interleukin 12 Receptor Subunit Beta 1) encodes a type I transmembrane protein that binds to interleukin-12 (IL12) and is involved in IL12 transduction. Mutations in *IL12RB1* damage the development of IL17-producing T lymphocytes and increase the susceptibility to Salmonella and mycobacterial infections (van de Vosse et al. 2013). Our PPI network analysis indicated that IL12RB1 and TYK2 had experimentally determined interactions, which were compiled from a set of public databases and were more likely to be credible (Q. C. Zhang et al. 2013). Gene *TYK2* had been previously identified to be associated with Covid-19 critical illness (Pairo-Castineira et al. 2021), implying the potential effects of *IL12RB1* to the aggravation of Covid-19. The gene *TRAF3IP3* (TRAF3 Interacting Protein 3) encodes a protein that plays essential roles in both innate and adaptive immunity. Knockout mouse experiments of this gene observed a decrease in white blood cell count in males and an increased susceptibility to bacterial infection (GARDIN and WHITE 2011). In our results, TRAF3IP3 was experimentally determined with protein TRAF3 encoded by gene *TRAF3*, which was included in a newly created pathway “Activation of NLRP3 inflammasome by SARS-CoV-2” (WP4876) (Siu et al. 2019). In response to viral infection, *TRAF3IP3* bridges *TRAF3* and *MAVS* leading to interferon production, indicating its probably strong relationship with Covid-19 disease.

**Figure 3.**
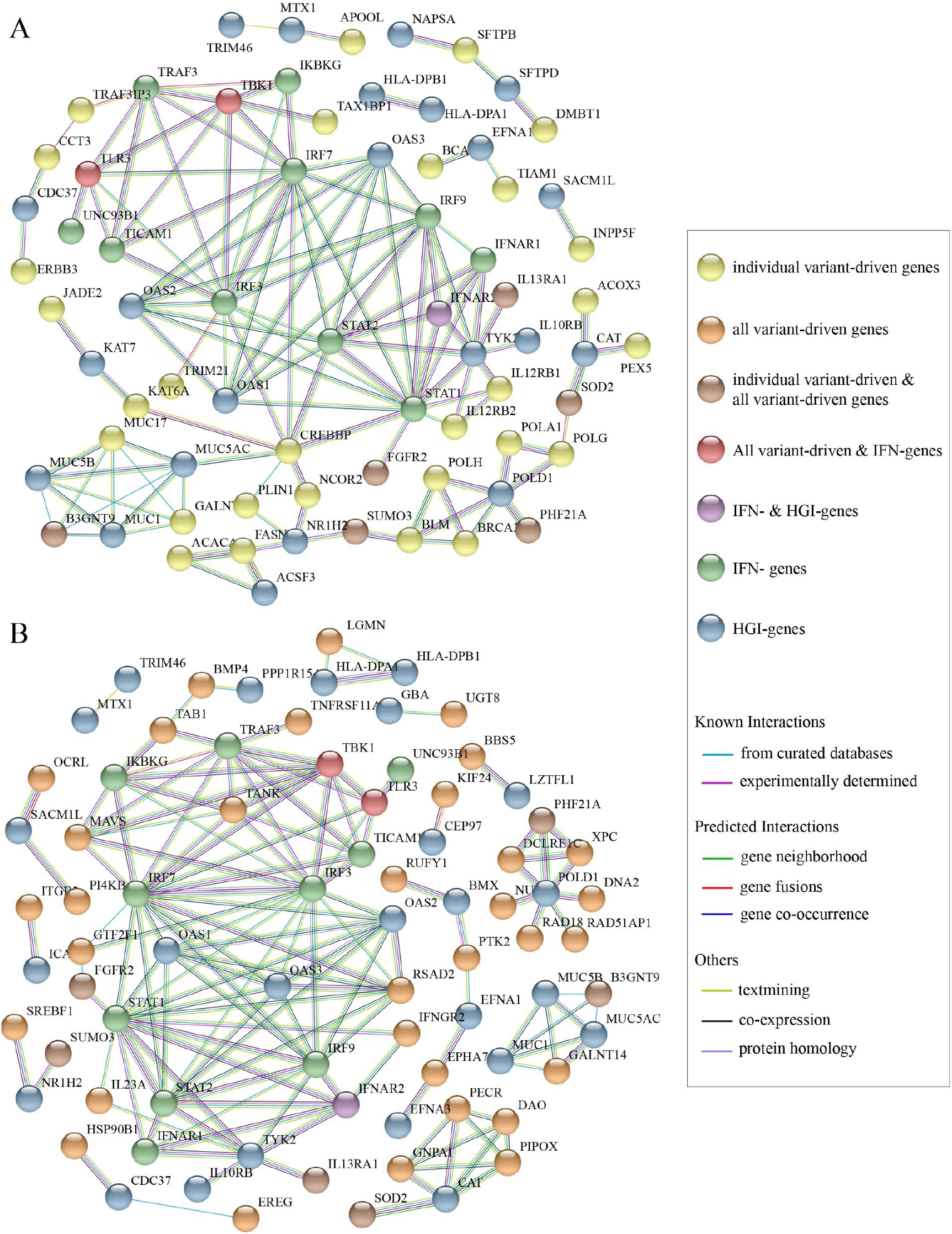
The results of protein-protein interaction network analysis. The plot of PPI network (A) between the 700 “individual variant-driven” genes with the 148 known genes. (B) between the 778 “all variant-driven” genes with the 148 known genes.

We also performed PPI network analysis for the 778 “all variant-driven” genes with the 148 known genes (Figure 3B), from which two genes, *TBK1* and *TLR3*, were highlighted. Specifically, *TBK1* (TANK Binding Kinase 1) encodes a protein that plays important roles in antiviral innate immune response and in regulating inflammatory response to foreign agents (Fitzgerald et al. 2003; Mori et al. 2004). A previous study observed colocalization of *TBK1* with the M protein of SARS-CoV-2, which might hinder the dsRNA-induced IFN production at the step or upstream of *TBK1* (Zheng et al. 2020). The gene *TLR3* (Toll Like Receptor 3) encodes a member of TLR family that plays a primary role in recognition of pathogen and innate immunity activation. It recognizes dsRNA participated in multiple viral infections and induces type I IFNs production (Kawai and Akira 2007).

In summary, our mutation accumulation analyses and PPI network analyses suggested that locus 3p21.31, *IL12RB1, TRAF3IP3, TBK1*, and *TLR3* as key regions in *severe* Covid-19 patients compared with *non-severe*, implying their functions and associations with Covid-19 severity.

## Discussion

SARS-CoV-2 is a strain of coronavirus and is highly pathogenic and transmissible. After exposure to the virus, ordinary people may do not develop noticeable symptoms or develop mild to moderate symptoms, while people with inborn errors of immunity tend to suffer severe and critical symptoms, or even death (S.-Y. Zhang et al. 2020). There is increasing evidence that the host genetic variants in genes related to immunodeficiency or inflammasomes might attribute to Covid-19 clinical manifestations (Elhabyan et al. 2020). A bunch of clinical drug treatments for Covid-19 were cultivated from this finding, including type I IFNs (e.g., IFN-α1b), TNF inhibitors, anti-IFN-γantibodies, JAK1 inhibitors, and STAT1 inhibitors (Ku, Chen, and Lai 2021).

In this work, we carried out the first study of rare variants in inborn errors of immunity genes associated with Covid-19 severity in Chinese population. The identified functional candidate pathways Tuberculosis, Primary Immunodeficiency, and Influenza A were previously known to be part of antiviral immune responses and viral eradication, and we discovered their potential influences in Covid-19. We also suggested several putative genetic regions probably involved in susceptibility and severity of Covid-19, including locus 3p21.31, genes *IL12RB1, TRAF3IP3, TBK1*, and *TLR3*.

Despite the many compelling and significant findings of our work, there are still a few limitations to be noted. First, the sample size we used is relatively small, and the limited sample size limits the statistical power for identifying rare variants. More studies with large sample sizes are demanded to validate our results and uncover more candidate variants. Second, even though our work has suggested several candidate genes and pathways potentially related to Covid-19 severity, the true underlying genetic mechanisms of how they affect disease progression need to be explored by more persuasive experimental designs.

Covid-19 is assessed as a complex infectious disease and affected many risk factors. Symptoms of Covid-19 are highly variable, ranging from unnoticeable to severe and even death. The host genetic background is only partly responsible for the phenotypic heterogeneity. In recent years, multi-omics studies have proven a powerful and successful strategy to provide a broader perspective in understanding disease development and biological phenomena. Several multi-omics analyses of Covid-19 have been proposed to integrate multiple “omes” data to unravel disease mechanisms at multiple omics levels (Overmyer et al. 2021; Su et al. 2020; Montaldo et al. 2021; P. Wu, Chen, et al. 2021; Stephenson et al. 2021). The integrative analyses of rare genome and other “omes” data (e.g., proteome, transcriptome, epigenome, metabolome, and microbiome) may inspire us to discover new risk factors for severe Covid-19 disease.

## Data Availability

All data produced in the present work are contained in the manuscript

## Author Contribution

X.J., H.Z. conceived the study, designed the research program, and managed the project.

F.C., F.Z., Y.J. collected the samples.

Y.L. finished the laboratory processing and data acquisition.

P.L., H.Z. performed the statistical analyses.

M.F advised on statistical methods.

P.L., H.Z. wrote the manuscript.

All authors participated in revising the manuscript.

## Acknowledgement

We thank the families who participated in the study and made this research study possible.

## Data Availability

The original contributions presented in the study are included in the article/Supplementary Material. Further inquiries can be directed to the corresponding author.

## Figures

**Supplementary Figure S1. Gene-based tests**. (A). The QQ plots of gene-based association analyses between severe and non-severe patients for (A). 42,730 candidate rare variants; (B) 38,548 rare likely-deleterious missense variants, and (C) 4,182 high-confidence pLoF variants

## References

Babaha, Fateme, and Nima Rezaei. 2020. “Primary Immunodeficiency Diseases in COVID-19 Pandemic: A Predisposing or Protective Factor?” The American Journal of the Medical Sciences. https://doi.org/10.1016/j.amjms.2020.07.027.

Bibert, Stéphanie, Nicolas Guex, Joao Lourenco, Thomas Brahier, Matthaios Papadimitriou-Olivgeris, Lauro Damonti, Oriol Manuel, et al. 2021. “Transcriptomic Signature Differences Between SARS-CoV-2 and Influenza Virus Infected Patients.” Frontiers in Immunology. https://www.frontiersin.org/article/10.3389/fimmu.2021.666163.

Browning, Brian L, and Sharon R Browning. 2016. “Genotype Imputation with Millions of Reference Samples.” American Journal of Human Genetics 98 (1): 116–26. https://doi.org/10.1016/j.ajhg.2015.11.020.

Casto, Amanda M, and Marcus W Feldman. 2011. “Genome-Wide Association Study SNPs in the Human Genome Diversity Project Populations: Does Selection Affect Unlinked SNPs with Shared Trait Associations?” PLoS Genetics 7 (1): e1001266. https://doi.org/10.1371/journal.pgen.1001266.

Chan, Kelvin Y K, Johannes C Y Ching, MS Xu, Annie N Y Cheung, Shea-Ping Yip, Loretta Y C Yam, Sik-To Lai, et al. 2007. “Association of ICAM3 Genetic Variant with Severe Acute Respiratory Syndrome.” The Journal of Infectious Diseases 196 (2): 271–80. https://doi.org/10.1086/518892.

Chang, Christopher C, Carson C Chow, Laurent Cam Tellier, Shashaank Vattikuti, Shaun M Purcell, and James J Lee. 2015. “Second-Generation PLINK: Rising to the Challenge of Larger and Richer Datasets.” GigaScience 4: 7. https://doi.org/10.1186/s13742-015-0047-8.

Clarke, Laura, Xiangqun Zheng-Bradley, Richard Smith, Eugene Kulesha, Chunlin Xiao, Iliana Toneva, Brendan Vaughan, et al. 2012. “The 1000 Genomes Project: Data Management and Community Access.” Nature Methods 9 (5): 459–62. https://doi.org/10.1038/nmeth.1974.

Cummings, Matthew J, Matthew R Baldwin, Darryl Abrams, Samuel D Jacobson, Benjamin J Meyer, Elizabeth M Balough, Justin G Aaron, et al. 2020. “Epidemiology, Clinical Course, and Outcomes of Critically Ill Adults with COVID-19 in New York City: A Prospective Cohort Study.” Lancet (London, England) 395 (10239): 1763–70. https://doi.org/10.1016/S0140-6736(20)31189-2.

Danecek, Petr, James K Bonfield, Jennifer Liddle, John Marshall, Valeriu Ohan, Martin O Pollard, Andrew Whitwham, et al. 2021. “Twelve Years of SAMtools and BCFtools.” GigaScience 10 (2): giab008. https://doi.org/10.1093/gigascience/giab008.

DePristo, Mark A, Eric Banks, Ryan Poplin, Kiran v Garimella, Jared R Maguire, Christopher Hartl, Anthony A Philippakis, et al. 2011. “A Framework for Variation Discovery and Genotyping Using Next-Generation DNA Sequencing Data.” Nature Genetics 43 (5): 491–98. https://doi.org/10.1038/ng.806.

Elhabyan, Abdelazeem, Saja Elyaacoub, Ehab Sanad, Abdelwahab Abukhadra, Asmaa Elhabyan, and Valentin Dinu. 2020. “The Role of Host Genetics in Susceptibility to Severe Viral Infections in Humans and Insights into Host Genetics of Severe COVID-19: A Systematic Review.” Virus Research 289 (November): 198163. https://doi.org/10.1016/j.virusres.2020.198163.

Ellinghaus, David, Frauke Degenhardt, Luis Bujanda, Maria Buti, Agustín Albillos, Pietro Invernizzi, Javier Fernández, et al. 2020. “Genomewide Association Study of Severe Covid-19 with Respiratory Failure.” The New England Journal of Medicine, June. https://doi.org/10.1056/NEJMoa2020283.

Fitzgerald, Katherine A, Sarah M McWhirter, Kerrie L Faia, Daniel C Rowe, Eicke Latz, Douglas T Golenbock, Anthony J Coyle, Sha-Mei Liao, and Tom Maniatis. 2003. “IKKepsilon and TBK1 Are Essential Components of the IRF3 Signaling Pathway.” Nature Immunology 4 (5): 491–96. https://doi.org/10.1038/ni921.

Freed, Donald, Rafael Aldana, Jessica Weber, and Jeremy Edwards. 2017. “The Sentieon Genomics Tools - A Fast and Accurate Solution to Variant Calling from next-Generation Sequence Data.” bioRxiv. https://doi.org/10.1101/115717.

Gardin, A, and J White. 2011. “The Sanger Mouse Genetics Programme: High Throughput Characterisation of Knockout Mice.” Acta Ophthalmologica 89 (s248): 0. https://doi.org/10.1111/j.1755-3768.2011.4451.x.

Gorbalenya, Alexander E, Susan C Baker, Ralph S Baric, Raoul J de Groot, Christian Drosten, Anastasia A Gulyaeva, Bart L Haagmans, et al. 2020. “The Species Severe Acute Respiratory Syndrome-Related Coronavirus: Classifying 2019-NCoV and Naming It SARS-CoV-2.” Nature Microbiology 5 (4): 536–44. https://doi.org/10.1038/s41564-020-0695-z.

Gorlov, I P, O Y Gorlova, M L Frazier, M R Spitz, and C I Amos. 2011. “Evolutionary Evidence of the Effect of Rare Variants on Disease Etiology.” Clinical Genetics 79 (3): 199–206. https://doi.org/10.1111/j.1399-0004.2010.01535.x.

Group, The TB/COVID-19 Global Study. 2021. “Tuberculosis and COVID-19 Co-Infection: Description of the Global Cohort.” European Respiratory Journal, January, 2102538. https://doi.org/10.1183/13993003.02538-2021.

Ho, Hsi-En, Sheryl Mathew, Michael J Peluso, and Charlotte Cunningham-Rundles. 2021. “Clinical Outcomes and Features of COVID-19 in Patients with Primary Immunodeficiencies in New York City.” The Journal of Allergy and Clinical Immunology. In Practice 9 (1): 490–493.e2. https://doi.org/10.1016/j.jaip.2020.09.052.

Hunt, Karen A, Vanisha Mistry, Nicholas A Bockett, Tariq Ahmad, Maria Ban, Jonathan N Barker, Jeffrey C Barrett, et al. 2013. “Negligible Impact of Rare Autoimmune-Locus Coding-Region Variants on Missing Heritability.” Nature 498 (7453): 232– 35. https://doi.org/10.1038/nature12170.

Initiative, COVID-19 Host Genetics, and Andrea Ganna. 2021. “Mapping the Human Genetic Architecture of COVID-19: An Update.” MedRxiv, January, 2021.11.08.21265944. https://doi.org/10.1101/2021.11.08.21265944.

Itan, Yuval, Lei Shang, Bertrand Boisson, Michael J Ciancanelli, Janet G Markle, Ruben Martinez-Barricarte, Eric Scott, et al. 2016. “The Mutation Significance Cutoff: Gene-Level Thresholds for Variant Predictions.” Nature Methods 13 (2): 109–10. https://doi.org/10.1038/nmeth.3739.

Kanehisa, Minoru, and Susumu Goto. 2000. “KEGG: Kyoto Encyclopedia of Genes and Genomes.” Nucleic Acids Research 28 (1): 27–30. https://doi.org/10.1093/nar/28.1.27.

Karczewski, Konrad J, Laurent C Francioli, Grace Tiao, Beryl B Cummings, Jessica Alföldi, Qingbo Wang, Ryan L Collins, et al. 2020. “The Mutational Constraint Spectrum Quantified from Variation in 141,456 Humans.” Nature 581 (7809): 434–43. https://doi.org/10.1038/s41586-020-2308-7.

Kawai, Taro, and Shizuo Akira. 2007. “Signaling to NF-KappaB by Toll-like Receptors.” Trends in Molecular Medicine 13 (11): 460–69. https://doi.org/10.1016/j.molmed.2007.09.002.

Kircher, Martin, Daniela M Witten, Preti Jain, Brian J O’Roak, Gregory M Cooper, and Jay Shendure. 2014. “A General Framework for Estimating the Relative Pathogenicity of Human Genetic Variants.” Nature Genetics 46 (3): 310–15. https://doi.org/10.1038/ng.2892.

Ku, Cheng-Lung, I-Ting Chen, and Ming-Zong Lai. 2021. “Infection-Induced Inflammation from Specific Inborn Errors of Immunity to COVID-19.” The FEBS Journal 288 (17): 5021–41. https://doi.org/10.1111/febs.15961.

Lee, Seunggeun, Michael C Wu, and Xihong Lin. 2012. “Optimal Tests for Rare Variant Effects in Sequencing Association Studies.” Biostatistics (Oxford, England) 13 (4): 762–75. https://doi.org/10.1093/biostatistics/kxs014.

Li, Miaoxin, Jiang Li, Mulin Jun Li, Zhicheng Pan, Jacob Shujui Hsu, Dajiang J Liu, Xiaowei Zhan, Junwen Wang, Youqiang Song, and Pak Chung Sham. 2017. “Robust and Rapid Algorithms Facilitate Large-Scale Whole Genome Sequencing Downstream Analysis in an Integrative Framework.” Nucleic Acids Research 45 (9): e75. https://doi.org/10.1093/nar/gkx019.

Mantovani, Stefania, Sergio Daga, Chiara Fallerini, Margherita Baldassarri, Elisa Benetti, Nicola Picchiotti, Francesca Fava, et al. 2021. “Rare Variants in Toll-like Receptor 7 Results in Functional Impairment and Downregulation of Cytokine-Mediated Signaling in COVID-19 Patients.” Genes and Immunity, December, 1– 6. https://doi.org/10.1038/s41435-021-00157-1.

Masood, Kiran Iqbal, Maliha Yameen, Javeria Ashraf, Saba Shahid, Syed Faisal Mahmood, Asghar Nasir, Nosheen Nasir, et al. 2021. “Upregulated Type I Interferon Responses in Asymptomatic COVID-19 Infection Are Associated with Improved Clinical Outcome.” Scientific Reports 11 (1): 22958. https://doi.org/10.1038/s41598-021-02489-4.

McLaren, William, Laurent Gil, Sarah E Hunt, Harpreet Singh Riat, Graham R S Ritchie, Anja Thormann, Paul Flicek, and Fiona Cunningham. 2016. “The Ensembl Variant Effect Predictor.” Genome Biology 17 (1): 122. https://doi.org/10.1186/s13059-016-0974-4.

Misawa, Kazuharu, Takanori Hasegawa, Eikan Mishima, Promsuk Jutabha, Motoshi Ouchi, Kaname Kojima, Yosuke Kawai, Masafumi Matsuo, Naohiko Anzai, and Masao Nagasaki. 2020. “Contribution of Rare Variants of the SLC22A12 Gene to the Missing Heritability of Serum Urate Levels.” Genetics 214 (4): 1079–90. https://doi.org/10.1534/genetics.119.303006.

Montaldo, Chiara, Francesco Messina, Isabella Abbate, Manuela Antonioli, Veronica Bordoni, Alessandra Aiello, Fabiola Ciccosanti, et al. 2021. “Multi-Omics Approach to COVID-19: A Domain-Based Literature Review.” Journal of Translational Medicine 19 (1): 501. https://doi.org/10.1186/s12967-021-03168-8.

Mori, Mitsuaki, Mitsutoshi Yoneyama, Takashi Ito, Kiyohiro Takahashi, Fuyuhiko Inagaki, and Takashi Fujita. 2004. “Identification of Ser-386 of Interferon Regulatory Factor 3 as Critical Target for Inducible Phosphorylation That Determines Activation.” The Journal of Biological Chemistry 279 (11): 9698–9702. https://doi.org/10.1074/jbc.M310616200.

Overmyer, Katherine A, Evgenia Shishkova, Ian J Miller, Joseph Balnis, Matthew N Bernstein, Trenton M Peters-Clarke, Jesse G Meyer, et al. 2021. “Large-Scale Multi-Omic Analysis of COVID-19 Severity.” Cell Systems 12 (1): 23–40.e7. https://doi.org/10.1016/j.cels.2020.10.003.

Pairo-Castineira, Erola, Sara Clohisey, Lucija Klaric, Andrew D Bretherick, Konrad Rawlik, Dorota Pasko, Susan Walker, et al. 2021. “Genetic Mechanisms of Critical Illness in COVID-19.” Nature 591 (7848): 92–98. https://doi.org/10.1038/s41586-020-03065-y.

Shelton, Janie F, Anjali J Shastri, Chelsea Ye, Catherine H Weldon, Teresa Filshtein-Sonmez, Daniella Coker, Antony Symons, Jorge Esparza-Gordillo, Stella Aslibekyan, and Adam Auton. 2021. “Trans-Ancestry Analysis Reveals Genetic and Nongenetic Associations with COVID-19 Susceptibility and Severity.” Nature Genetics 53 (6): 801–8. https://doi.org/10.1038/s41588-021-00854-7.

Sherry, S T, M.-H. Ward, M Kholodov, J Baker, L Phan, E M Smigielski, and K Sirotkin. 2001. “DbSNP: The NCBI Database of Genetic Variation.” Nucleic Acids Research 29 (1): 308–11. https://doi.org/10.1093/nar/29.1.308.

Shields, Adrian M, Siobhan O Burns, Sinisa Savic, and Alex G Richter. 2021. “COVID-19 in Patients with Primary and Secondary Immunodeficiency: The United Kingdom Experience.” The Journal of Allergy and Clinical Immunology 147 (3): 870–875.e1. https://doi.org/10.1016/j.jaci.2020.12.620.

Siu, Kam-Leung, Kit-San Yuen, Carlos Castaño-Rodriguez, Zi-Wei Ye, Man-Lung Yeung, Sin-Yee Fung, Shuofeng Yuan, et al. 2019. “Severe Acute Respiratory Syndrome Coronavirus ORF3a Protein Activates the NLRP3 Inflammasome by Promoting TRAF3-Dependent Ubiquitination of ASC.” FASEB Journal : Official Publication of the Federation of American Societies for Experimental Biology 33 (8): 8865–77. https://doi.org/10.1096/fj.201802418R.

Smieszek, Sandra P, Vasilios M Polymeropoulos, Changfu Xiao, Christos M Polymeropoulos, and Mihael H Polymeropoulos. 2021. “Loss-of-Function Mutations in IFNAR2 in COVID-19 Severe Infection Susceptibility.” Journal of Global Antimicrobial Resistance 26 (July): 239–40. https://doi.org/10.1016/j.jgar.2021.06.005.

Stephenson, Emily, Gary Reynolds, Rachel A Botting, Fernando J Calero-Nieto, Michael D Morgan, Zewen Kelvin Tuong, Karsten Bach, et al. 2021. “Single-Cell Multi-Omics Analysis of the Immune Response in COVID-19.” Nature Medicine 27 (5): 904–16. https://doi.org/10.1038/s41591-021-01329-2.

Su, Yapeng, Daniel Chen, Dan Yuan, Christopher Lausted, Jongchan Choi, Chengzhen L Dai, Valentin Voillet, et al. 2020. “Multi-Omics Resolves a Sharp Disease-State Shift between Mild and Moderate COVID-19.” Cell 183 (6): 1479–1495.e20. https://doi.org/10.1016/j.cell.2020.10.037.

Sweeney, Timothy E, Shane Lofgren, Purvesh Khatri, and Angela J Rogers. 2017. “Gene Expression Analysis to Assess the Relevance of Rodent Models to Human Lung Injury.” American Journal of Respiratory Cell and Molecular Biology 57 (2): 184–92. https://doi.org/10.1165/rcmb.2016-0395OC.

Szklarczyk, Damian, Annika L Gable, David Lyon, Alexander Junge, Stefan Wyder, Jaime Huerta-Cepas, Milan Simonovic, et al. 2019. “STRING V11: Protein– Protein Association Networks with Increased Coverage, Supporting Functional Discovery in Genome-Wide Experimental Datasets.” Nucleic Acids Research 47 (D1): D607–13. https://doi.org/10.1093/nar/gky1131.

Vosse, Esther van de, Margje H Haverkamp, Noe Ramirez-Alejo, Mónica Martinez-Gallo, Lizbeth Blancas-Galicia, Ayşe Metin, Ben Zion Garty, et al. 2013. “IL-12Rβ1 Deficiency: Mutation Update and Description of the IL12RB1 Variation Database.” Human Mutation 34 (10): 1329–39. https://doi.org/10.1002/humu.22380.

Wang, Fang, Shujia Huang, Huirong Gao, Yuwen Zhou, Changxiang Lai, Zhichao Li, Wenjie Xian, et al. 2020. “Initial Whole Genome Sequencing and Analysis of the Host Genetic Contribution to COVID-19 Severity and Susceptibility.” MedRxiv, January, 2020.06.09.20126607. https://doi.org/10.1101/2020.06.09.20126607.

Williams, Frances M K, Maxim Freydin, Massimo Mangino, Simon Couvreur, Alessia Visconti, Ruth C E Bowyer, Caroline I le Roy, et al. 2020. “Self-Reported Symptoms of Covid-19 Including Symptoms Most Predictive of SARS-CoV-2 Infection, Are Heritable.” MedRxiv, January, 2020.04.22.20072124. https://doi.org/10.1101/2020.04.22.20072124.

Wu, Michael C, Peter Kraft, Michael P Epstein, Deanne M Taylor, Stephen J Chanock, David J Hunter, and Xihong Lin. 2010. “Powerful SNP-Set Analysis for Case-Control Genome-Wide Association Studies.” American Journal of Human Genetics 86 (6): 929–42. https://doi.org/10.1016/j.ajhg.2010.05.002.

Wu, Peng, Dongsheng Chen, Wencheng Ding, Ping Wu, Hongyan Hou, Yong Bai, Yuwen Zhou, et al. 2021. “The Trans-Omics Landscape of COVID-19.” Nature Communications 12 (1): 4543. https://doi.org/10.1038/s41467-021-24482-1.

Wu, Peng, Lin Ding, Xiaodong Li, Siyang Liu, Fanjun Cheng, Qing He, Mingzhong Xiao, et al. 2021. “Trans-Ethnic Genome-Wide Association Study of Severe COVID-19.” Communications Biology 4 (1): 1034. https://doi.org/10.1038/s42003-021-02549-5.

Wu, Zunyou, and Jennifer M McGoogan. 2020. “Characteristics of and Important Lessons From the Coronavirus Disease 2019 (COVID-19) Outbreak in China: Summary of a Report of 72 314 Cases From the Chinese Center for Disease Control and Prevention.” JAMA 323 (13): 1239–42. https://doi.org/10.1001/jama.2020.2648.

Yang, Xiaobo, Yuan Yu, Jiqian Xu, Huaqing Shu, Jia’an Xia, Hong Liu, Yongran Wu, et al. 2020. “Clinical Course and Outcomes of Critically Ill Patients with SARS-CoV-2 Pneumonia in Wuhan, China: A Single-Centered, Retrospective, Observational Study.” The Lancet. Respiratory Medicine 8 (5): 475–81. https://doi.org/10.1016/S2213-2600(20)30079-5.

Zhang, Fan, Matthew Flickinger, Sarah A Gagliano Taliun, Gonçalo R Abecasis, Laura J Scott, Steven A McCaroll, Carlos N Pato, Michael Boehnke, and Hyun Min Kang. 2020. “Ancestry-Agnostic Estimation of DNA Sample Contamination from Sequence Reads.” Genome Research 30 (2): 185–94. https://doi.org/10.1101/gr.246934.118.

Zhang, Qian, Zhiyong Liu, Marcela Moncada-Velez, Jie Chen, Masato Ogishi, Benedetta Bigio, Rui Yang, et al. 2020. “Inborn Errors of Type I IFN Immunity in Patients with Life-Threatening COVID-19.” Science 370 (6515). https://doi.org/10.1126/science.abd4570.

Zhang, Qiangfeng Cliff, Donald Petrey, José Ignacio Garzón, Lei Deng, and Barry Honig. 2013. “PrePPI: A Structure-Informed Database of Protein-Protein Interactions.” Nucleic Acids Research 41 (Database issue): D828–33. https://doi.org/10.1093/nar/gks1231.

Zhang, Shen-Ying, Qian Zhang, Jean-Laurent Casanova, Helen C Su, and COVID Team. 2020. “Severe COVID-19 in the Young and Healthy: Monogenic Inborn Errors of Immunity?” Nature Reviews. Immunology 20 (8): 455–56. https://doi.org/10.1038/s41577-020-0373-7.

Zheng, Yi, Meng-Wei Zhuang, Lulu Han, Jing Zhang, Mei-Ling Nan, Peng Zhan, Dongwei Kang, Xinyong Liu, Chengjiang Gao, and Pei-Hui Wang. 2020. “Severe Acute Respiratory Syndrome Coronavirus 2 (SARS-CoV-2) Membrane (M) Protein Inhibits Type I and III Interferon Production by Targeting RIG-I/MDA-5 Signaling.” Signal Transduction and Targeted Therapy 5 (1): 299. https://doi.org/10.1038/s41392-020-00438-7.

Zhu, Huanhuan, Fang Zheng, Linxuan Li, Yan Jin, Yuxue Luo, Zhen Li, JingYu Zeng, et al. 2021. “A Chinese Host Genetic Study Discovered Type I Interferons and Causality of Cholesterol Levels and WBC Counts on COVID-19 Severity.” MedRxiv, January, 2021.06.04.21258335. https://doi.org/10.1101/2021.06.04.21258335.

Zhu, Na, Dingyu Zhang, Wenling Wang, Xingwang Li, Bo Yang, Jingdong Song, Xiang Zhao, et al. 2020. “A Novel Coronavirus from Patients with Pneumonia in China, 2019.” The New England Journal of Medicine 382 (8): 727–33. https://doi.org/10.1056/NEJMoa2001017.

Zuk, Or, Stephen F Schaffner, Kaitlin Samocha, Ron Do, Eliana Hechter, Sekar Kathiresan, Mark J Daly, Benjamin M Neale, Shamil R Sunyaev, and Eric S Lander. 2014. “Searching for Missing Heritability: Designing Rare Variant Association Studies.” Proceedings of the National Academy of Sciences 111 (4): E455 LP–E464. https://doi.org/10.1073/pnas.1322563111.

